# Projected Aging Among People with HIV in the United States: A Modeling Analysis in 24 States

**DOI:** 10.64898/2026.01.30.26345234

**Authors:** Andrew Zalesak, Parastu Kasaie, Zoe Dansky, Keri N. Althoff, David W. Dowdy, Maunank Shah, Anthony T. Fojo, Melissa Schnure

**Author notes:** **Corresponding Author**: Andrew Zalesak.

## Abstract

**Importance:** As the population living with HIV in the US ages, state-level projections of the aging dynamics among people with diagnosed HIV (PWDH) will be needed to inform local planning and intervention efforts.

**Objective:** We sought to explore how aging dynamics of the population with HIV in the US are expected to differ at the state level between 2025 and 2040.

**Design, Setting, and Participants:** We projected epidemic trajectories from 2025 to 2040 in 24 US states comprising 86% of PWDH in the US using a calibrated model of HIV transmission.

**Main Outcomes and Measures:** We estimated change in median age of PWDH over age 13, from 2025 to 2040, for each state.

**Results:** We project that by 2040, the median age of adult PWDH in the 24 states will rise from 51 to 62 years, and over half of adult PWDH will be over the age of 65. Our projections suggest substantial heterogeneities in age distributions by state. More populous and urban states with higher median ages of PWDH in 2025 are projected to experience even further aging of the population with diagnosed HIV in the coming 15 years. By contrast, more rural and less populous states tend to have younger-aged HIV epidemics that were not projected to age substantially over time.

**Conclusions and Relevance:** Although the overall population of persons with diagnosed HIV in the US is projected to age substantially, these effects will unfold differently across states. In the coming years, healthcare systems will need to plan to adapt to changing state-level demographic patterns among PWDH.

**Key Points:** *Question:* How will the age distribution of people living with diagnosed HIV change between 2025 and 2040?

*Findings:* Using a calibrated model of HIV transmission in 24 US states, we project that by 2040, the median age of adult PWDH in the 24 states will rise from 51 to 62 years, and over half of adult PWDH will be over the age of 65. Our projections suggest substantial differences in age distribution by state.

*Meaning:* In the coming years, federal and local healthcare planning will need to adapt to changing state-level demographic patterns among PWDH.

## Introduction

The United States (US) has made considerable progress in curtailing its HIV epidemic over the past several decades.^1,2^. The widespread availability of effective antiretroviral therapy (ART) has increased the life expectancy of the 1.2 million people with diagnosed HIV (PWDH) in the US^2^. By the end of 2022, over half of all individuals with diagnosed HIV in the US were aged 50 years or older^3^.

People aging with HIV have complex and multifaceted healthcare needs that extend beyond viral suppression. Aging PWDH are at increased risk for age-related conditions including cardiovascular, metabolic, and renal disease, and certain non-AIDS-defining cancers^4^. They are also at greater risk of developing multiple comorbidities^5^. As the population living with HIV ages, local healthcare systems – as well as the Medicare, Medicaid, and Ryan White programs – will need to manage the increasing burden of age-related comorbidities among this population.

Mathematical models of infectious disease contribute to our understanding of the aging dynamics of local HIV epidemics. While existing studies explore aging among PWDH in the US, they have typically been conducted at the national level, focus only on certain risk groups such as men who have sex with men (MSM), or do not project beyond 2030^6-9^. The objective of our study was to explore how aging dynamics of the population with HIV are expected to differ at the state level in the US between 2025 and 2040.

## Methods

### Model Structure and Calibration

The Johns Hopkins Epidemiologic and Economic Model (JHEEM) is a dynamic, compartmental model of HIV transmission in the US, stratifying the adult population by age, race/ethnicity, sex, HIV acquisition risk factors, and HIV status^10^ (**Supplemental Figure S1)**. In this analysis, we model HIV epidemics in 24 states comprising 86% of PWDH in the US: Alabama, Arkansas, Arizona, California, Colorado, Florida, Georgia, Illinois, Kentucky, Louisiana, Maryland, Michigan, Missouri, Mississippi, North Carolina, New York, Ohio, Oklahoma, South Carolina, Tennessee, Texas, Virginia, Washington, and Wisconsin. These states were chosen to represent varied geographic regions and prioritization within the Ending the HIV Epidemic initiative^11^.

The model calibration process at the state level followed a similar Bayesian methodology to previously-published models^12^. This approach supports robust estimation of key parameters by aligning model outputs with calibration targets for population demographics (general population size, general population mortality, immigration, and emigration), HIV dynamics (new diagnoses, diagnosed prevalence, and mortality among PWDH), and HIV prevention and care (e.g., use of pre-exposure prophylaxis, HIV testing, awareness of HIV status, and viral suppression – see Supplement).

### Modeled Scenario and Outcomes

After calibration to available data from 1970 to 2024, we projected the models forward from 2025 to 2040 across the 24 states. Our projections followed a “status quo” scenario, assuming that recent trends in HIV transmission dynamics and public health programming will continue into the future. This baseline scenario also captures temporary disruptions in the care cascade due to the COVID pandemic from 2020 to 2022.

Our primary outcome was the change in median age of PWDH over age 13, from 2025 and 2040. Secondary outcomes included the median age of PWDH in 2025 and 2040; the state-level number and percentage of PWDH who were aged 55 or older (as a percentage of all adults aged 13+ with diagnosed HIV in the state) in 2025 and 2040; and the number and percentage of PWDH aged 65+ (the threshold for Medicare eligibility for most people^13^) in 2025 and 2040. For all outcomes, we report mean values and corresponding 95% credible intervals (2.5^th^ and 97.5^th^ percentiles) across 1,000 simulations. We also present results by subgroup, specifically HIV acquisition risk (MSM vs non-MSM) and race/ethnicity (Hispanic and Black, and other non-Black, non-Hispanic PWDH).

The JHEEM represents five age brackets: 13-24, 25-34, 35-44, 45-54, and 55+ years old, reflecting the age stratifications in US Centers for Disease Control and Prevention (CDC) surveillance data^14^. In order to calculate the median age and the percentage of PWDH over 65, we used a Penalized Composite Link Model (PCLM) smoothing function to derive single-year age estimates from the binned age projections, from the R package *ungroup*^15,16^.

### Secondary Analysis

To explore which factors may drive differences between states, we examined the correlations between five key quantities and our outcomes (change in median age of adult PWDH and change in the percentage of PWDH who are aged 55+): (1) the number of new diagnoses as a percentage of diagnosed prevalence in 2025; (2) the median age of adult PWDH in 2025; (3) the median age of new HIV diagnoses in 2025; (4) prevalence of diagnosed HIV as a percentage of general population in 2025; and (5) the relative urbanicity of a state’s HIV epidemic, calculated as the average 2020 US-census urbanicity of each county in a state, weighted by the number of PWDH in each county in 2021. We calculated Spearman correlation coefficients between our outcomes (the average change in median age and the average change in the percentage of PWDH who are aged 55+) and the average value of each potential determinant of aging across all 1,000 simulations in each state. We visualized these relationships with scatterplots.

### Sensitivity Analysis

We conducted sensitivity analyses to identify the parameters that had the strongest influence on the change in median age between 2025 and 2040. For each state, we calculated partial rank correlation coefficients (PRCC, a measure of the correlation between each parameter and the outcome, adjusted for all other parameters). For highly influential parameters, we visualized the outcome in the 20% of simulations with the highest values of the parameter, contrasting to the 20% of simulations with the lowest values^17^.

## Results

### Model Calibration and Total Prevalence

Overall, calibrated simulations reproduced our epidemiologic targets well – see **Figure 1** for the example state of California and www.jheem.org/aging for other states. Aggregated across all 24 states, the model projected the total number of PWDH to rise from 917,115 (95% credible interval: 911,107 to 922,885) in 2025 to 1,001,144 (972,173 to 1,025,696) in 2040 (**Figure 2**, “Total”). Prevalence of diagnosed HIV was projected to increase in all states except for California, Illinois, Maryland, Michigan, and New York, where the model projected slight decreases in diagnosed prevalence during this period (**Figure 2**). Most states showed a bimodal age distribution, where ages 55+ and 35-44 were the largest age categories (**Figure 2**).

**Figure 1:**
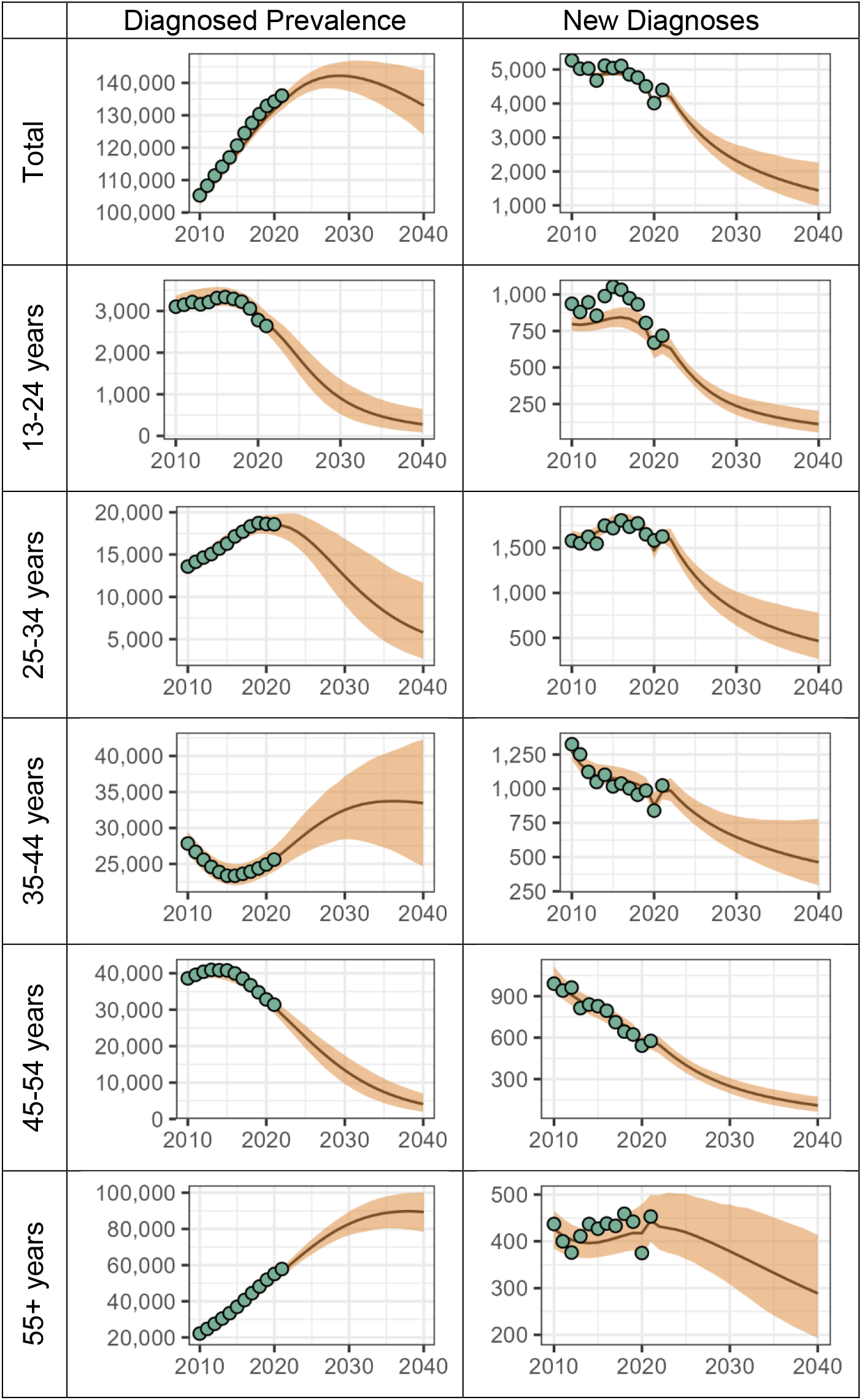
Projected Diagnosed Prevalence and New Diagnoses of HIV in California by Age. Black lines denote the mean value across 1,000 model simulations for number of people living with diagnosed HIV (left-hand panels) and new diagnoses of HIV (right-hand panels). The orange-shaded ribbons denote the 95% credible intervals shown. Green dots indicate CDC surveillance data.

**Figure 2:**
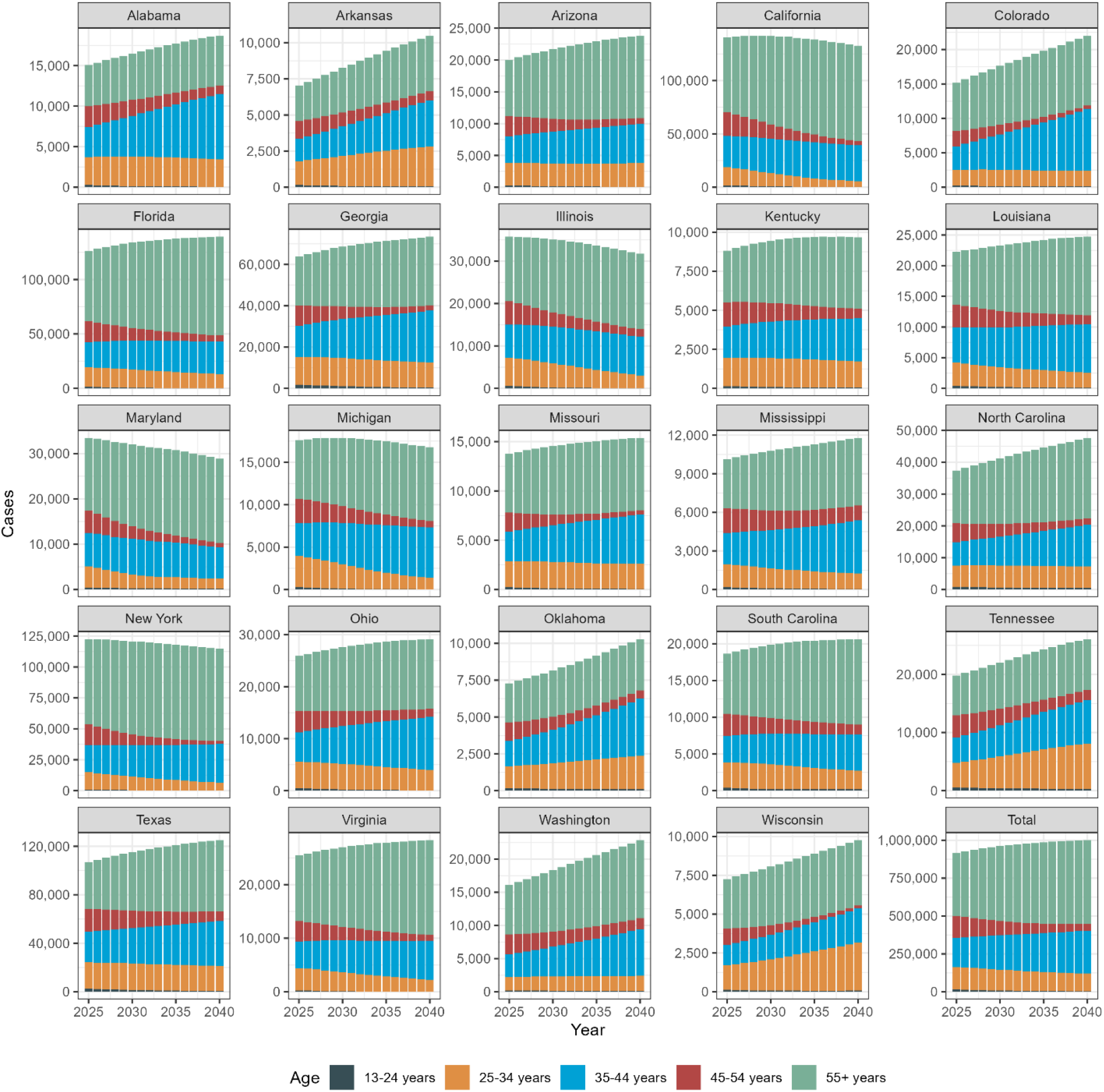
Projected Age Distribution of People with Diagnosed HIV Over Time. Plots show model projections of the number of adults living with diagnosed HIV from 2025 and 2040 by state and for the total among the 24 modeled states. The height of the bars is proportional to the number of individuals, stratified by age – from 13–24-year-olds at the bottom to 55+ year-olds at the top.

### Median Age Among Adults with Diagnosed HIV

In the 15 years from 2025 to 2040, the median age among adults with diagnosed HIV was projected to shift 11 years older across all states, from 51 years (51 to 52) to 62 years (60 to 65, **Figure 3**). States with large, urban, and older-aged epidemics in 2025 had the greatest projected increases in median age. For example, California’s median age increased by 18 (15 to 20) years, rising from 53 (52 to 56) in 2025 to 71 (68 to 74) in 2040; Maryland, New York, and Florida had similar profiles. Meanwhile, states with small, rural, and younger-aged epidemics were projected to see decreasing median ages, with generally more uncertainty in their model projections. Arkansas’ median age among adult PWDH was projected to decrease by 6 (−8 to −1) years, from 45 (44 to 47) in 2025 to 39 (37 to 46) in 2040; Alabama, Mississippi, Oklahoma, and Tennessee had similar profiles.

**Figure 3:**
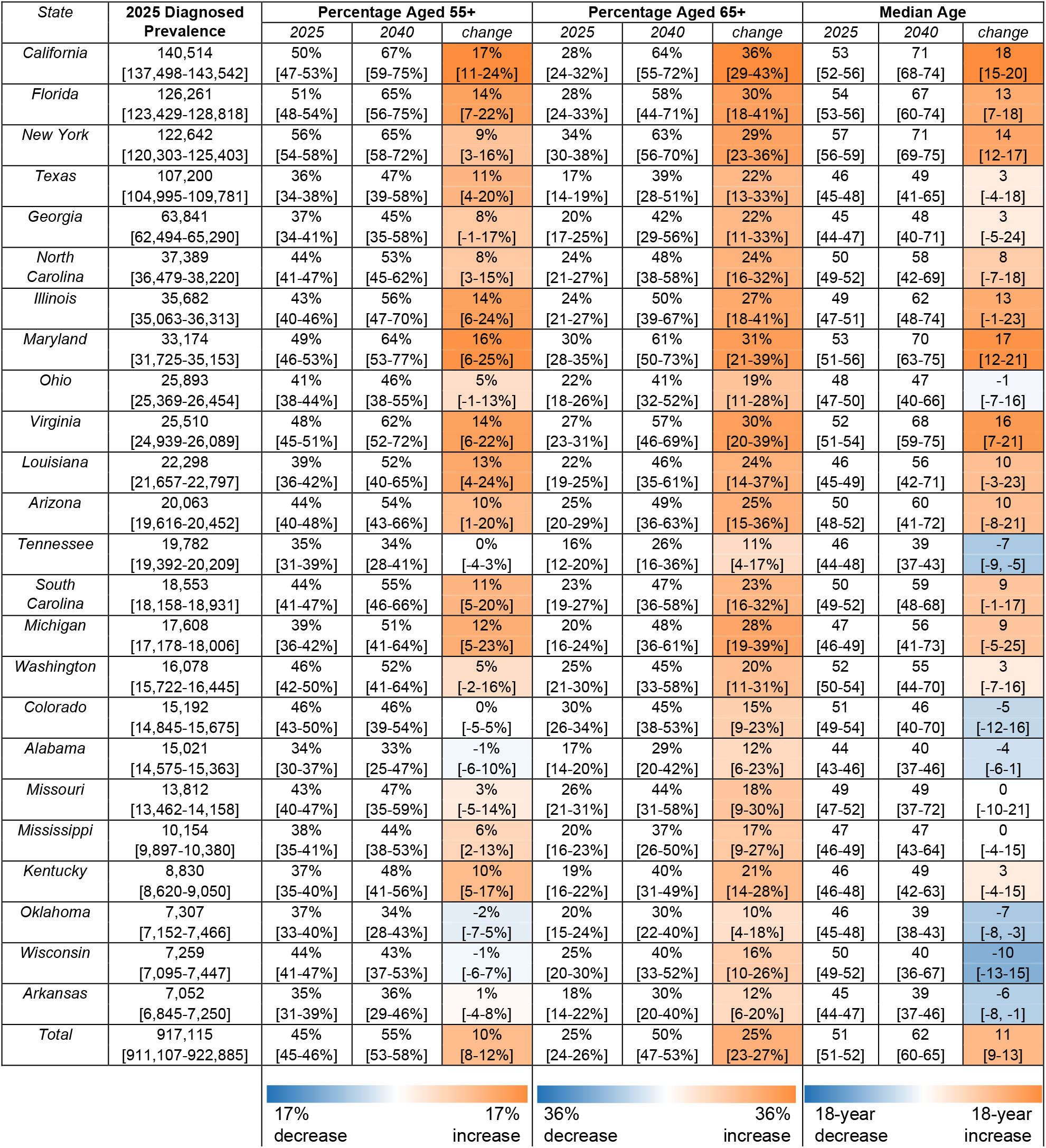
Projected Aging Among People with Diagnosed HIV by State. Values are the mean model projections and 95% credible intervals across 1,000 simulations. States are ordered by the 2025 prevalence of diagnosed HIV among all adults over age 13. Percentage aged 55+ and 65+ indicate the percentage of all diagnosed adults (over age 13) living with HIV who fall into these age categories, with values for 2025, 2040, and the change between these two years. The median age is for all adults with diagnosed HIV (over age 13), with values for 2025, 2040 and the change between these two years. Cells are shaded according to the change between years within each measure, with darker orange values indicating states with greater aging and darker blue values indicating states with increasingly younger populations.

### Median Age Change by Subgroup

The median age of adult MSM with diagnosed HIV across the 24 states was projected to increase by 11 (−3 to 19) years, rising from 48 (47 to 49) in 2025 to 59 (45 to 67) in 2040 (**Supplemental Figure S2)**. The median age for non-MSM PWDH was projected to increase by 8 (5 to 9) years, rising from 54 (54 to 55) in 2025 to 62 (60 to 64) in 2040.

Among our three modeled racial/ethnic categories, Hispanic and Black PWDH were substantially younger in 2025 than other (non-Black, non-Hispanic) PWDH (**Supplemental Figure S3**). All three groups were projected to age, with non-Black, non-Hispanic PWDH showing the most pronounced rate of aging. Specifically, median age was projected to rise from 48 (48 to 49) in 2025 to 55 (47 to 63) in 2040 among non-Hispanic Black PWDH, from 48 (47 to 50) to 63 (52 to 69) among Hispanic PWDH, and from 56 (56 to 58) to 67 (66 to 69) among other PWDH.

### Secondary Outcomes: Changes in People with HIV Aged 55+ and 65+

The number of PWDH aged 55+ was projected to increase from 417,091 (410,673 to 424,924) in 2025 to 552,810 (533,818 to 575,511) in 2040, rising by 10 percentage points (8 to 12), from 45% (45 to 46%) in 2025 to 55% (53 to 58%) in 2040 (**Figure 3** and **Supplemental Figure S4**). States with larger baseline percentages of PWDH aged 55+ in 2025 tended to have greater increases in this percentage by 2040. For example, California’s percentage of PWDH aged 55+ rose by 17 (11 to 24) percentage points from 50% (47 to 53%) in 2025 to 67% (59 to 75%) in 2040, while Arkansas’s percentage stayed at approximately one-third, shifting only slightly from 35% (31 to 39%) in 2025 to 36% (29 to 46%) in 2040.

The absolute number of PWDH across all 24 states aged 65+ was projected to increase by 280,541 (256,228 to 302,594) individuals, rising from 206,379 (198,967 to 214,600) in 2025 to 486,920 (460,288 to 513,112) in 2040 (**Figure 3** and **Supplemental Figure S5**). The percentage of all PWDH aged 65+ rose by 25% (23 to 27%) from 25% (24 to 26%) in 2025 to 50% (47 to 53%) in 2040. State-level patterns roughly mirrored those in the aged 55+ results above.

### Secondary Analysis of State-Level Variation

Among our five candidate state-level determinants of change in median age, the covariate with the highest correlation (Spearman R: −0.73) was the ratio of new diagnoses to diagnosed prevalence in 2025, which approximates the average transmission rate in our model. In states with higher transmission, aging of existing cases was offset by a relatively large number of younger individuals newly diagnosed with HIV. Other determinants had a lower correlation with change in median age (see **Figure 4**). Correlations with the change in percentage of PWDH who are aged 55 or older were weaker but qualitatively similar (**Supplemental Figure S5**).

**Figure 4:**
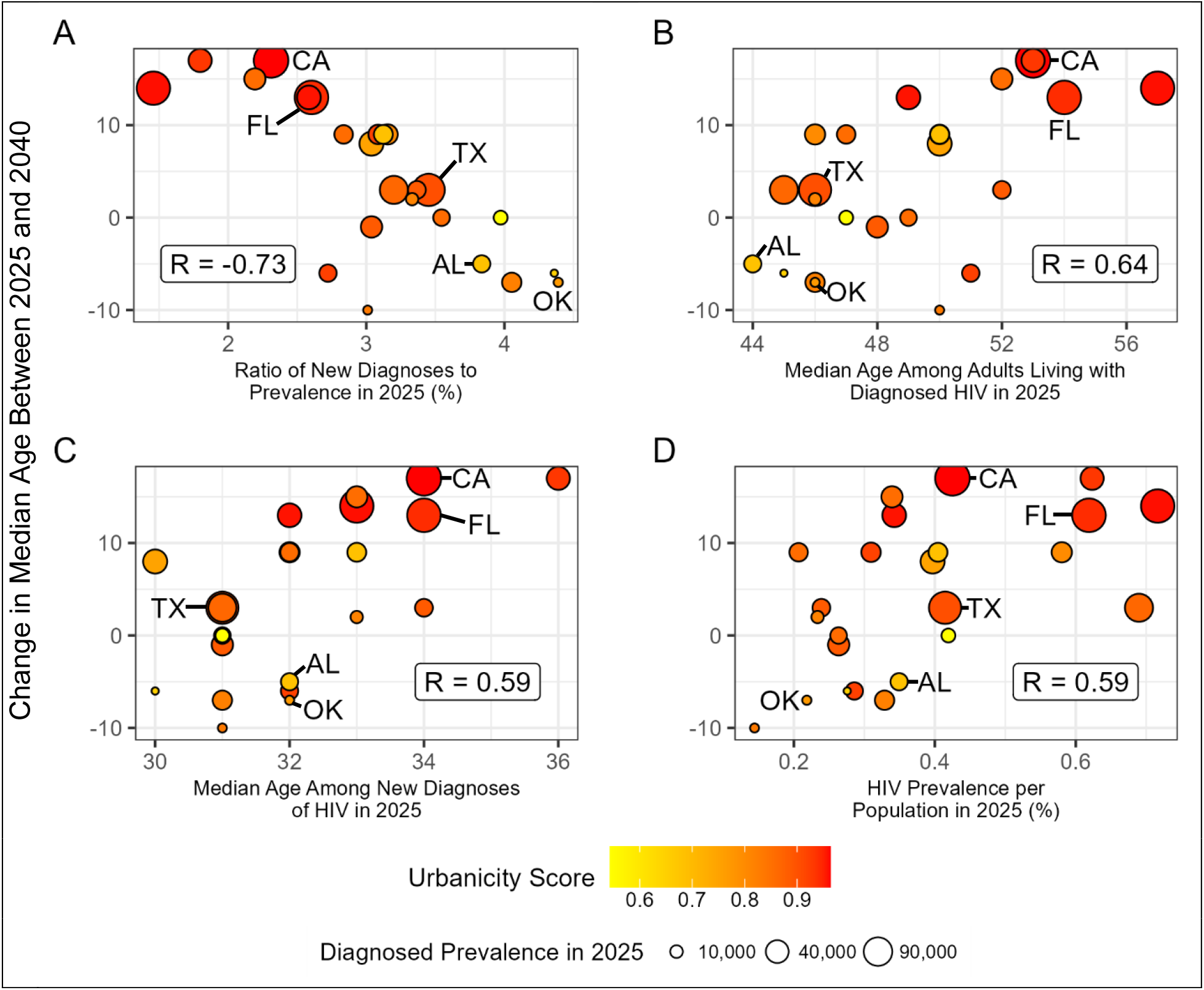
State-level Characteristics v. Projected Change in Median Age of People with Diagnosed HIV. Scatterplots show the projected change in median age of adult PWDH versus four state-level characteristics, averaged across 1,000 simulations in each state: (A) the ratio of new HIV diagnoses to HIV prevalence in 2025; (B) the median age among adults living with HIV in 2025; (C) the median age among new HIV diagnoses in 2025; and (D) prevalence as percentage of the general population in 2025. Each point represents one of the 24 states; points are sized by that state’s diagnosed prevalence in 2025 and are shaded by the urbanicity of the state’s HIV epidemic, which has a Spearman correlation of 0.54 with the projected change in median age. Five states are highlighted: Alabama (“AL”), California (“CA”), Florida (“FL”), Oklahoma (“OK”), and Texas (“TX”). Each panel is labeled with the Spearman correlation coefficient.

In general, states with large, urban epidemics with a higher median age in 2025 – such as California, New York, Florida, and Maryland – were projected to have larger increases in median age from 2025 to 2040 (see **Figure 4**). Conversely, states with small, rural epidemics with a lower median age in 2025 – such as Alabama, Arkansas, Mississippi, Oklahoma, and Tennessee – tended to see small rises or even decreases in the median age. Several states were exceptions to this trend: for instance, Georgia and Texas aged less than states with similarly large and urban epidemics, but their projections aligned with states with similar ratios of new diagnoses to diagnosed prevalence.

### Sensitivity Analyses

**Supplemental Figure S6** shows a sensitivity analysis of model parameters most strongly associated with the change in median age among adult PWDH, on average across all cities. The most influential parameter was the percentage of 35-44-year-old MSM PWDH who are 44 years old in 2020, with an average partial rank correlation coefficient of 0.215. This parameter governs the rate at which 35-44-year-olds are expected to age into the 45-54-year-old bracket. Across all 24 states, the 20% of simulations with the highest percentage of 44-year-olds in that age bracket in 2020 were projected to have an increase of 13 years in median age (Interquartile range: 12 to 14), compared to 8 years (IQR: 6 to 10) in the 20% of simulations with the smallest percentage. The next two most influential parameters governed the age distribution within the 35-44-year-old age bracket among MSM PWDH in 2010 and within the 45-44-year-old age bracket among MSM PWDH in 2010.

## Discussion

We used a calibrated model of HIV transmission to project the future age distribution of PWDH in 24 US states. By 2040, the median age of adult PWDH across these states will rise from 51 to 62 years, and over half of adult PWDH will be over the age of 65. Our projections showed substantial differences in these age distributions by state. More populous and urban states with higher current median ages of PWDH were projected to experience even further aging of the population with diagnosed HIV in the coming 15 years. By contrast, more rural and less populous states tended to have younger HIV epidemics. This contrast can be seen between California and Oklahoma: while we project the median age of adult PWDH in California will reach 67 by 2040 (with two-thirds of PWDH over the age of 55), the median age of adult PWDH in Oklahoma will be only 41 by 2040, and only one-third of PWDH will be over the age of 55.

As PWDH age across the country, federal and local HIV programming will need to adapt to support their evolving needs, especially in light of growing workforce shortages in HIV care^18^. Our projections suggest a substantial increase in number of PWDH aged 55+ (by over 135,000) and those aged 65+ (by over 280,000) by 2040. This will translate to a higher demand for Medicare and other federally funded programs to provide comprehensive care and support services – for example, among the Medicare-eligible population (aged 65+) the number of PWDH is projected to grow from 5.5 per thousand in 2025 to 6.8 per thousand in 2040. Federal programs can support this change by providing funding for integrated HIV and primary care, developing national guidelines for age-appropriate HIV care (including comorbidity management), supporting workforce training in geriatric HIV care, expanding coverage for age-related conditions and investing in future research and programs that promote healthy aging and reduce long term health care costs.

At a local level, the percentage increase in number of aging PWDH can vary substantially, with some states experiencing an increase over 30% in the coming decade. Meeting this demand will require a rapid shift in the structure and scope of local HIV programs. Of note, local programs can integrate HIV and primary care, ensuring that HIV clinicians are prepared to manage age-associated comorbidities among their patients, including cardiovascular and metabolic diseases. They should also support mental and cognitive health by offering counseling, cognitive screening and services tailored to aging population. Further social and community services – such as transportation, housing, and peer support – can enhance quality of life, reduce isolation and promote healthy aging among older PWDH.

Other methods have been used to project the future age distribution of people living with HIV in the US. Althoff and colleagues used an agent-based simulation model of HIV care in the US, projecting a modest increase in overall median age among ART users to 52 years by 2030^6^. This is similar to our projections, although the study represented the epidemic at the national level and did not dynamically model transmission as we did. A study by the CDC used a life-table-based cohort-component model of PWDH in the US, projecting an overall median age among PWDH of 55 years by 2040 in a “status quo” scenario assuming an annual percent change in new diagnoses of −3%^19^. Our model projected a higher median age of 62 by 2040 while showing a similar change in new diagnoses across the model period. This difference may again be due to how transmission is represented in our model: by dynamically modeling transmission, new diagnoses begin to occur among older individuals as the existing population with HIV ages. This shift, along with our slightly lower projected mortality rates among PWDH, likely contributes to an older projected population with HIV compared to methodologies that do not explicitly model transmission.

There are several limitations to this study. First, we assume that there will be no major changes to HIV care, prevention, and testing services during the modeled period. While the absolute number of PWDH aged 55+ is less sensitive to changes in transmission, if reductions to HIV care and prevention result in higher transmission rates, there will be more infections than we project here, and a lower percentage of PWDH will be over 55 years old; consequently, the median age will be younger than we projected. Second, we assume that age-specific mortality rates observed today will continue largely unchanged into the future; if advances in medicine or healthcare delivery extend the lifespans of people with HIV, we would expect an older population with HIV than projected here. Third, we assume that people from different regions within a state are uniformly likely to form transmission linkages. If regions within a state do not mix uniformly and have very different age distributions, then we may under- or over-estimate the age of new infections.

Our modeling approach has several advantages. Using a Bayesian calibration method allows us to quantify the uncertainty around our model parameters and calibration data and incorporate it into our projections. By modeling at the state, rather than the national, level, we can capture the dynamics of local epidemics. Finally, local policymakers can explore projections for individual states and demographic subgroups with our web tool, available at www.jheem.org/aging.

Using a local-level model of HIV transmission in 24 states, we project that the population of people diagnosed with HIV in the US will age substantially in the coming years. These effects will be felt differently across states: more populous, urban states with older populations with HIV are likely to age more than less populous, more rural states with younger populations with HIV. These projections can inform planning for managing and preventing age-related comorbidities to optimize the health of people aging with HIV.

## Supporting information

Technical Supplement

## Data Availability

All data produced are available online at www.jheem.org/aging.

