## Supplementary material for "Projected Aging Among People with HIV in the United States: A Modeling Analysis in 24 States": Technical Supplement

### Model Structure

#### Supplemental Figure S1: Johns Hopkins Epidemiological and Economic Model Structure

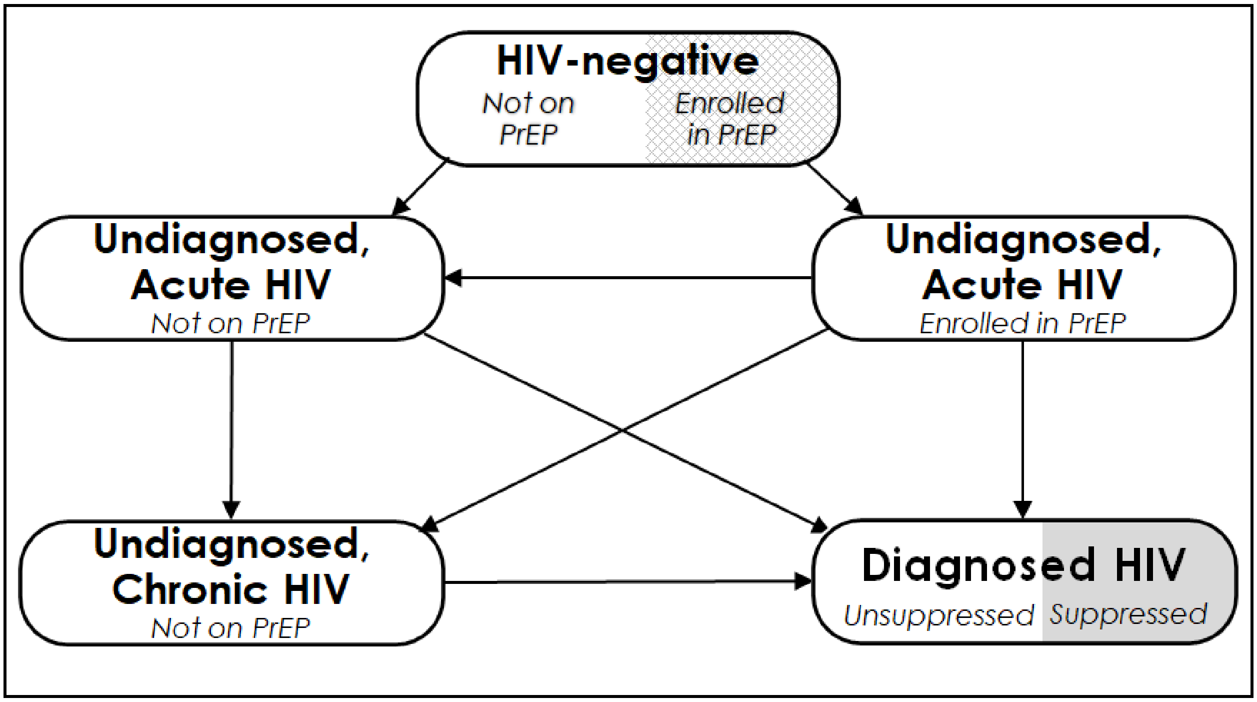

This figure depicts the compartments representing HIV status. Each of the five compartments is further stratified by age (13–24, 25–34, 35–44, 45–54, and ≥55 years), race/ethnicity (Black, Hispanic, and other), sex and sexual behavior (female, heterosexual male, and men who have sex with men (MSM), and intravenous drug use history (never used, active use, and prior use). “Acute HIV” refers to the first 2.9 months following infection, during which risk of transmission is high.

### Additional Results

#### Supplemental Figure S2: Projected Median Age of Adults with Diagnosed HIV by Race and State

| *State* | **2025 Diagnosed Prevalence** | **Black** | | | **Hispanic** | | | **Other** | | |
|---|---|---|---|---|---|---|---|---|---|---|
| | | *2025* | *2040* | *change* | *2025* | *2040* | *change* | *2025* | *2040* | *change* |
| *California* | 140,514 [137,498-143,542] | 50 [48-53] | 57 [42-76] | 7 [-8-24] | 47 [45-49] | 66 [44-76] | 19 [-1-28] | 63 [60-66] | 77 [72-81] | 14 [10-17] |
| *Florida* | 126,261 [123,429-128,818] | 52 [50-55] | 65 [51-75] | 13 [1-21] | 52 [50-57] | 68 [58-78] | 16 [7-24] | 60 [58-64] | 71 [63-78] | 11 [4-16] |
| *New York* | 122,642 [120,303-125,403] | 57 [54-61] | 69 [53-77] | 12 [-1-19] | 59 [56-64] | 82 [78-86] | 23 [18-26] | 60 [56-66] | 76 [66-83] | 16 [9-22] |
| *Texas* | 107,200 [104,995-109,781] | 44 [42-46] | 48 [39-72] | 4 [-4-27] | 43 [41-46] | 46 [39-66] | 3 [-3-21] | 55 [52-59] | 70 [43-80] | 15 [-8-23] |
| *Georgia* | 63,841 [62,494-65,290] | 43 [42-45] | 46 [39-76] | 3 [-4-31] | 43 [41-46] | 61 [40-85] | 18 [-2-41] | 55 [52-60] | 57 [39-79] | 2 [-15-22] |
| *North Carolina* | 37,389 [36,479-38,220] | 49 [47-52] | 64 [42-76] | 15 [-6-25] | 43 [42-45] | 40 [37-44] | -3 [-5 to 1] | 55 [51-58] | 64 [41-73] | 9 [-10-17] |
| *Illinois* | 35,682 [35,063-36,313] | 43 [42-46] | 55 [39-83] | 12 [-4-38] | 48 [46-52] | 75 [64-83] | 27 [18-34] | 57 [51-64] | 72 [53-82] | 15 [0-22] |
| *Maryland* | 33,174 [31,725-35,153] | 53 [51-62] | 77 [72-83] | 24 [18-28] | 46 [44-53] | 57 [41-83] | 11 [-3-33] | 52 [50-55] | 42 [41-44] | -10 [-12 to 8] |
| *Ohio* | 25,893 [25,369-26,454] | 43 [41-46] | 41 [37-50] | -2 [-5-5] | 45 [43-49] | 54 [40-80] | 9 [-4-33] | 53 [50-57] | 65 [42-77] | 12 [-8-22] |
| *Virginia* | 25,510 [24,939-26,089] | 50 [48-53] | 76 [63-85] | 26 [15-35] | 43 [42-46] | 56 [40-90] | 13 [-3-46] | 59 [56-67] | 71 [60-80] | 12 [4-18] |
| *Louisiana* | 22,298 [21,657-22,797] | 45 [44-47] | 58 [43-76] | 13 [-1-29] | 44 [42-49] | 57 [39-78] | 13 [-3-31] | 52 [47-57] | 52 [40-74] | 0 [-11-19] |
| *Arizona* | 20,063 [19,616-20,452] | 44 [41-47] | 51 [37-80] | 7 [-5-34] | 44 [42-48] | 56 [39-76] | 12 [-4-30] | 57 [54-61] | 65 [49-74] | 8 [-5-16] |
| *Tennessee* | 19,782 [19,392-20,209] | 41 [40-43] | 37 [34-40] | -4 [-7 to 2] | 42 [41-45] | 45 [38-78] | 3 [-3-34] | 51 [49-55] | 50 [37-65] | -1 [-13-11] |
| *South Carolina* | 18,553 [18,158-18,931] | 49 [46-52] | 62 [48-73] | 13 [2-23] | 45 [43-47] | 49 [42-74] | 4 [-2-28] | 53 [51-56] | 49 [43-59] | -4 [-9-3] |
| *Michigan* | 17,608 [17,178-18,006] | 43 [42-46] | 52 [40-77] | 9 [-2-32] | 44 [42-47] | 54 [39-87] | 10 [-4-42] | 53 [50-59] | 70 [42-82] | 17 [-9-26] |
| *Washington* | 16,078 [15,722-16,445] | 46 [45-49] | 43 [40-55] | -3 [-7-6] | 44 [42-47] | 42 [37-60] | -2 [-6-14] | 57 [54-61] | 73 [64-79] | 16 [10-21] |
| *Colorado* | 15,192 [14,845-15,675] | 45 [44-49] | 43 [37-77] | -2 [-8-29] | 42 [41-44] | 48 [39-86] | 6 [-3-43] | 60 [54-69] | 53 [41-76] | -7 [-22-12] |
| *Alabama* | 15,021 [14,575-15,363] | 41 [39-44] | 39 [36-52] | -2 [-4-9] | 47 [45-51] | 64 [43-82] | 17 [-2-34] | 50 [49-53] | 41 [39-44] | -9 [-11 to 7] |
| *Missouri* | 13,812 [13,462-14,158] | 44 [42-48] | 54 [37-79] | 10 [-6-33] | 44 [42-48] | 38 [35-45] | -6 [-9 to 2] | 55 [51-59] | 50 [37-73] | -5 [-16-16] |
| *Mississippi* | 10,154 [9,897-10,380] | 47 [46-48] | 53 [46-71] | 6 [0-23] | 45 [42-49] | 61 [40-87] | 16 [-3-41] | 47 [45-51] | 39 [37-42] | -8 [-11 to 5] |
| *Kentucky* | 8,830 [8,620-9,050] | 42 [40-45] | 42 [36-67] | 0 [-6-23] | 42 [41-46] | 56 [41-79] | 14 [-1-35] | 48 [47-51] | 59 [41-72] | 11 [-6-23] |
| *Oklahoma* | 7,307 [7,152-7,466] | 40 [38-43] | 40 [36-65] | 0 [-3-23] | 41 [38-44] | 39 [34-62] | -2 [-5-18] | 49 [47-53] | 40 [37-49] | -9 [-11 to 3] |
| *Wisconsin* | 7,259 [7,095-7,447] | 41 [39-45] | 35 [33-40] | -6 [-8 to 4] | 46 [45-50] | 50 [37-82] | 4 [-9-34] | 57 [54-62] | 57 [38-76] | 0 [-19-16] |
| *Arkansas* | 7,052 [6,845-7,250] | 40 [38-43] | 39 [35-48] | -1 [-4-5] | 41 [39-45] | 42 [36-68] | 1 [-4-24] | 49 [48-52] | 41 [37-58] | -8 [-12-6] |
| *Total* | 917,115 [911,107-922,885] | 48 [48-49] | 55 [47-63] | 7 [-1-14] | 48 [47-50] | 63 [52-69] | 15 [5-19] | 56 [56-58] | 67 [66-69] | 11 [9-12] |

27-year decrease — 27-year increase

Values are the mean model projections and 95% credible intervals across 1,000 simulations. States are ordered by the 2025 prevalence of diagnosed HIV among all adults over age 13. Median age is for all adults with diagnosed HIV (over age 13), with values for 2025, 2040 and the change between these two years. Cells are shaded according to the change between years within each measure, with darker orange values indicating states with greater aging and darker blue values indicating states with increasingly younger populations.

#### Supplemental Figure S3: Projected Median Age of Adults with Diagnosed HIV by HIV Acquisition Risk Group and State

| *State* | **2025 Diagnosed Prevalence** | **MSM** | | | **Non-MSM** | | |
|---|---|---|---|---|---|---|---|
| | | *2025* | *2040* | *change* | *2025* | *2040* | *change* |
| *California* | 140,514 [137,498-143,542] | 53 [51-56] | 71 [66-77] | 18 [14 to 23] | 55 [53-59] | 69 [57-79] | 14 [4 to 22] |
| *Florida* | 126,261 [123,429-128,818] | 52 [50-56] | 74 [68-80] | 22 [16 to 27] | 56 [54-59] | 66 [56-75] | 10 [2 to 17] |
| *New York* | 122,642 [120,303-125,403] | 50 [48-54] | 71 [41-84] | 21 [-7 to 32] | 63 [60-66] | 73 [65-80] | 10 [4 to 16] |
| *Texas* | 107,200 [104,995-109,781] | 43 [42-45] | 43 [37-70] | 0 [-5 to 25] | 49 [48-52] | 61 [52-75] | 12 [3 to 24] |
| *Georgia* | 63,841 [62,494-65,290] | 41 [40-43] | 39 [37-44] | -2 [-4 to 2] | 52 [49-58] | 65 [45-86] | 13 [-5 to 31] |
| *North Carolina* | 37,389 [36,479-38,220] | 43 [42-46] | 41 [37-66] | -2 [-6 to 20] | 56 [55-59] | 65 [59-72] | 9 [4 to 14] |
| *Illinois* | 35,682 [35,063-36,313] | 44 [42-47] | 67 [40-85] | 23 [-4 to 39] | 53 [51-56] | 61 [51-78] | 8 [0 to 24] |
| *Maryland* | 33,174 [31,725-35,153] | 45 [44-48] | 82 [76-86] | 37 [30 to 41] | 55 [53-59] | 64 [57-71] | 9 [3 to 15] |
| *Ohio* | 25,893 [25,369-26,454] | 45 [44-48] | 45 [38-76] | 0 [-7 to 29] | 50 [48-52] | 53 [44-61] | 3 [-5 to 10] |
| *Virginia* | 25,510 [24,939-26,089] | 47 [45-50] | 72 [40-88] | 25 [-5 to 40] | 57 [54-64] | 73 [59-85] | 16 [5 to 27] |
| *Louisiana* | 22,298 [21,657-22,797] | 43 [41-46] | 48 [37-74] | 5 [-4 to 29] | 49 [48-52] | 62 [48-77] | 13 [1 to 26] |
| *Arizona* | 20,063 [19,616-20,452] | 49 [47-52] | 59 [38-77] | 10 [-9 to 27] | 52 [50-55] | 58 [45-73] | 6 [-5 to 20] |
| *Tennessee* | 19,782 [19,392-20,209] | 43 [41-45] | 36 [34-39] | -7 [-8 to -5] | 49 [47-51] | 48 [41-56] | -1 [-6 to 6] |
| *South Carolina* | 18,553 [18,158-18,931] | 45 [43-47] | 64 [41-83] | 19 [-4 to 37] | 54 [54-56] | 59 [55-67] | 5 [1 to 12] |
| *Michigan* | 17,608 [17,178-18,006] | 43 [42-46] | 50 [38-79] | 7 [-4 to 34] | 52 [50-57] | 66 [52-84] | 14 [1 to 28] |
| *Washington* | 16,078 [15,722-16,445] | 51 [49-55] | 56 [41-72] | 5 [-10 to 19] | 51 [50-54] | 55 [46-70] | 4 [-4 to 16] |
| *Colorado* | 15,192 [14,845-15,675] | 48 [46-52] | 45 [39-77] | -3 [-10 to 27] | 51 [48-55] | 55 [43-76] | 4 [-7 to 22] |
| *Alabama* | 15,021 [14,575-15,363] | 41 [40-43] | 39 [36-46] | -2 [-5 to 3] | 49 [46-52] | 44 [37-55] | -5 [-11 to 4] |
| *Missouri* | 13,812 [13,462-14,158] | 47 [44-50] | 43 [35-71] | -4 [-10 to 22] | 51 [49-54] | 62 [45-78] | 11 [-4 to 25] |
| *Mississippi* | 10,154 [9,897-10,380] | 43 [42-46] | 45 [38-77] | 2 [-5 to 32] | 49 [48-52] | 50 [46-63] | 1 [-3 to 12] |
| *Kentucky* | 8,830 [8,620-9,050] | 45 [44-47] | 47 [39-68] | 2 [-6 to 22] | 47 [46-50] | 53 [42-65] | 6 [-4 to 17] |
| *Oklahoma* | 7,307 [7,152-7,466] | 44 [43-47] | 38 [36-42] | -6 [-9 to -4] | 46 [45-49] | 44 [40-54] | -2 [-6 to 5] |
| *Wisconsin* | 7,259 [7,095-7,447] | 46 [44-49] | 37 [35-45] | -9 [-12 to -3] | 54 [52-57] | 65 [52-81] | 11 [0 to 26] |
| *Arkansas* | 7,052 [6,845-7,250] | 42 [40-45] | 38 [35-49] | -4 [-7 to 4] | 49 [47-51] | 44 [41-52] | -5 [-7 to 1] |
| *Total* | 917,115 [911,107-922,885] | 48 [47-49] | 59 [45-67] | 11 [-3 to 19] | 54 [54-55] | 62 [60-64] | 8 [5 to 9] |

37-year decrease — 37-year increase

Values are the mean model projections and 95% credible intervals across 1,000 simulations. States are ordered by the 2025 prevalence of diagnosed HIV among all adults over age 13. Median age is for all adults with diagnosed HIV (over age 13), with values for 2025, 2040 and the change between these two years. Cells are shaded according to the change between years within each measure, with darker orange values indicating states with greater aging and darker blue values indicating states with increasingly younger populations. Results are shown for MSM (men who have sex with men) and non-MSM.

### Supplemental Figure S4: Diagnosed Prevalence Projections with Estimated Populations of People with Diagnosed HIV Aged 55+ and 65+

| *State* | 2025 Diagnosed Prevalence | Number Aged 55+ | | | Number Aged 65+ | | |
|---|---|---|---|---|---|---|---|
| | | *2025* | *2040* | *change* | *2025* | *2040* | *change* |
| *State* | 140,514 [137,498-143,542] | 70,024 [65,956-74,277] | 89,408 [78,383-100,109] | 19,384 [11,274-27,031] | 39,499 [33,686-45,380] | 85,701 [73,811-96,117] | 46,203 [36,438-54,780] |
| *California* | 126,261 [123,429-128,818] | 64,700 [61,181-68,566] | 91,496 [80,767-106,063] | 26,796 [18,299-37,780] | 35,733 [30,687-41,277] | 81,650 [64,757-100,163] | 45,917 [32,445-61,268] |
| *Florida* | 122,642 [120,303-125,403] | 68,852 [65,678-71,994] | 74,912 [66,824-84,789] | 6,060 [-676-14,310] | 41,634 [37,428-46,169] | 72,444 [64,609-81,748] | 30,811 [22,832-40,324] |
| *New York* | 107,200 [104,995-109,781] | 38,685 [35,915-41,236] | 59,400 [50,022-71,931] | 20,715 [13,258-31,171] | 17,875 [15,258-20,517] | 48,830 [36,760-63,388] | 30,955 [19,692-43,185] |
| *Texas* | 63,841 [62,494-65,290] | 23,875 [21,933-26,114] | 32,831 [25,816-39,222] | 8,957 [3,603-14,183] | 12,884 [10,837-15,560] | 30,394 [21,754-38,112] | 17,511 [10,404-23,995] |
| *Georgia* | 37,389 [36,479-38,220] | 16,568 [15,443-17,557] | 25,162 [21,030-29,270] | 8,595 [5,249-12,083] | 8,990 [7,663-10,188] | 22,879 [18,052-27,285] | 13,889 [10,066-17,466] |
| *North Carolina* | 35,682 [35,063-36,313] | 15,209 [14,206-16,345] | 17,807 [15,471-20,933] | 2,598 [516-4,859] | 8,428 [7,419-9,801] | 16,001 [13,097-19,970] | 7,573 [5,127-10,691] |
| *Illinois* | 33,174 [31,725-35,153] | 16,098 [15,121-17,547] | 18,292 [15,043-20,320] | 2,195 [-151-3,524] | 9,971 [9,205-11,426] | 17,356 [14,629-19,104] | 7,384 [5,038-8,889] |
| *Maryland* | 25,893 [25,369-26,454] | 10,553 [9,802-11,373] | 13,498 [11,414-16,201] | 2,945 [1,335-5,138] | 5,660 [4,754-6,641] | 12,007 [9,435-15,159] | 6,347 [4,216-8,928] |
| *Ohio* | 25,510 [24,939-26,089] | 12,253 [11,533-13,001] | 17,675 [15,174-20,266] | 5,423 [3,209-7,579] | 6,977 [5,984-8,025] | 16,350 [13,221-19,409] | 9,373 [6,578-11,868] |
| *Virginia* | 22,298 [21,657-22,797] | 8,637 [7,909-9,406] | 12,937 [10,189-16,089] | 4,299 [2,252-6,750] | 4,891 [4,157-5,658] | 11,604 [8,760-15,113] | 6,712 [4,351-9,687] |
| *Louisiana* | 20,063 [19,616-20,452] | 8,833 [8,098-9,544] | 13,118 [10,796-16,381] | 4,286 [2,419-7,033] | 4,919 [4,007-5,780] | 11,882 [8,923-15,676] | 6,962 [4,757-10,012] |
| *Arizona* | 19,782 [19,392-20,209] | 6,848 [6,143-7,593] | 8,759 [7,219-10,833] | 1,910 [1,037-3,472] | 3,066 [2,400-3,946] | 6,621 [4,403-8,616] | 3,555 [1,965-5,254] |
| *Tennessee* | 18,553 [18,158-18,931] | 8,115 [7,503-8,763] | 11,720 [9,802-15,446] | 3,605 [2,181-6,725] | 4,343 [3,596-5,088] | 9,989 [7,591-13,650] | 5,646 [3,941-8,797] |
| *South Carolina* | 17,608 [17,178-18,006] | 6,870 [6,239-7,485] | 8,692 [6,845-10,895] | 1,822 [553-3,484] | 3,607 [2,843-4,309] | 8,147 [5,951-10,358] | 4,540 [3,039-6,326] |
| *Michigan* | 16,078 [15,722-16,445] | 7,433 [6,792-8,037] | 11,793 [9,854-13,842] | 4,360 [2,876-5,989] | 4,052 [3,384-4,908] | 10,250 [8,002-12,501] | 6,198 [4,427-8,064] |
| *Washington* | 15,192 [14,845-15,675] | 7,049 [6,548-7,647] | 10,247 [8,767-12,178] | 3,198 [2,044-4,685] | 4,493 [3,874-5,237] | 9,983 [8,560-11,909] | 5,491 [4,074-7,217] |
| *Colorado* | 15,021 [14,575-15,363] | 5,045 [4,570-5,474] | 6,150 [4,767-8,168] | 1,105 [47-2,886] | 2,532 [2,099-3,024] | 5,375 [3,928-7,279] | 2,843 [1,691-4,511] |
| *Alabama* | 13,812 [13,462-14,158] | 5,964 [5,473-6,456] | 7,366 [6,015-8,672] | 1,402 [429-2,381] | 3,596 [2,938-4,264] | 6,999 [5,290-8,459] | 3,403 [2,159-4,503] |
| *Missouri* | 10,154 [9,897-10,380] | 3,853 [3,532-4,172] | 5,292 [4,325-6,461] | 1,439 [744-2,342] | 2,007 [1,665-2,375] | 4,377 [3,098-5,978] | 2,370 [1,329-3,722] |
| *Mississippi* | 8,830 [8,620-9,050] | 3,300 [3,057-3,532] | 4,620 [3,994-5,407] | 1,320 [839-1,943] | 1,697 [1,400-1,967] | 3,934 [3,052-4,776] | 2,237 [1,557-2,929] |
| *Kentucky* | 7,307 [7,152-7,466] | 2,669 [2,413-2,956] | 3,538 [2,956-4,338] | 869 [368-1,496] | 1,438 [1,120-1,766] | 3,102 [2,359-4,058] | 1,663 [1,037-2,407] |
| *Oklahoma* | 7,259 [7,095-7,447] | 3,185 [2,976-3,409] | 4,221 [3,700-4,774] | 1,036 [664-1,461] | 1,799 [1,470-2,151] | 3,939 [3,129-4,648] | 2,140 [1,547-2,732] |
| *Wisconsin* | 7,052 [6,845-7,250] | 2,474 [2,219-2,733] | 3,876 [3,080-4,853] | 1,402 [805-2,213] | 1,239 [971-1,548] | 3,166 [2,186-4,290] | 1,927 [1,149-2,862] |
| *Arkansas* | 917,115 [911,107-922,885] | 417,091 [410,673-424,924] | 552,810 [533,818-575,511] | 135,720 [120,067-153,442] | 206,379 [198,967-214,600] | 486,920 [460,288-513,112] | 280,541 [256,228-302,594] |

Values are the mean model projections and 95% credible intervals across 1,000 simulations. States are ordered by the 2025 prevalence of diagnosed HIV among all adults over age 13. Numbers aged 55+ and 65+ indicate the estimated number of diagnosed adults (over age 13) living with HIV who fall into these age categories, with values for 2025, 2040, and the change between these two years.

### Additional Secondary Analysis

#### Supplemental Figure S5: State-level Characteristics v. Projected Change in Percentage of People with Diagnosed HIV who are Aged 55 or Older

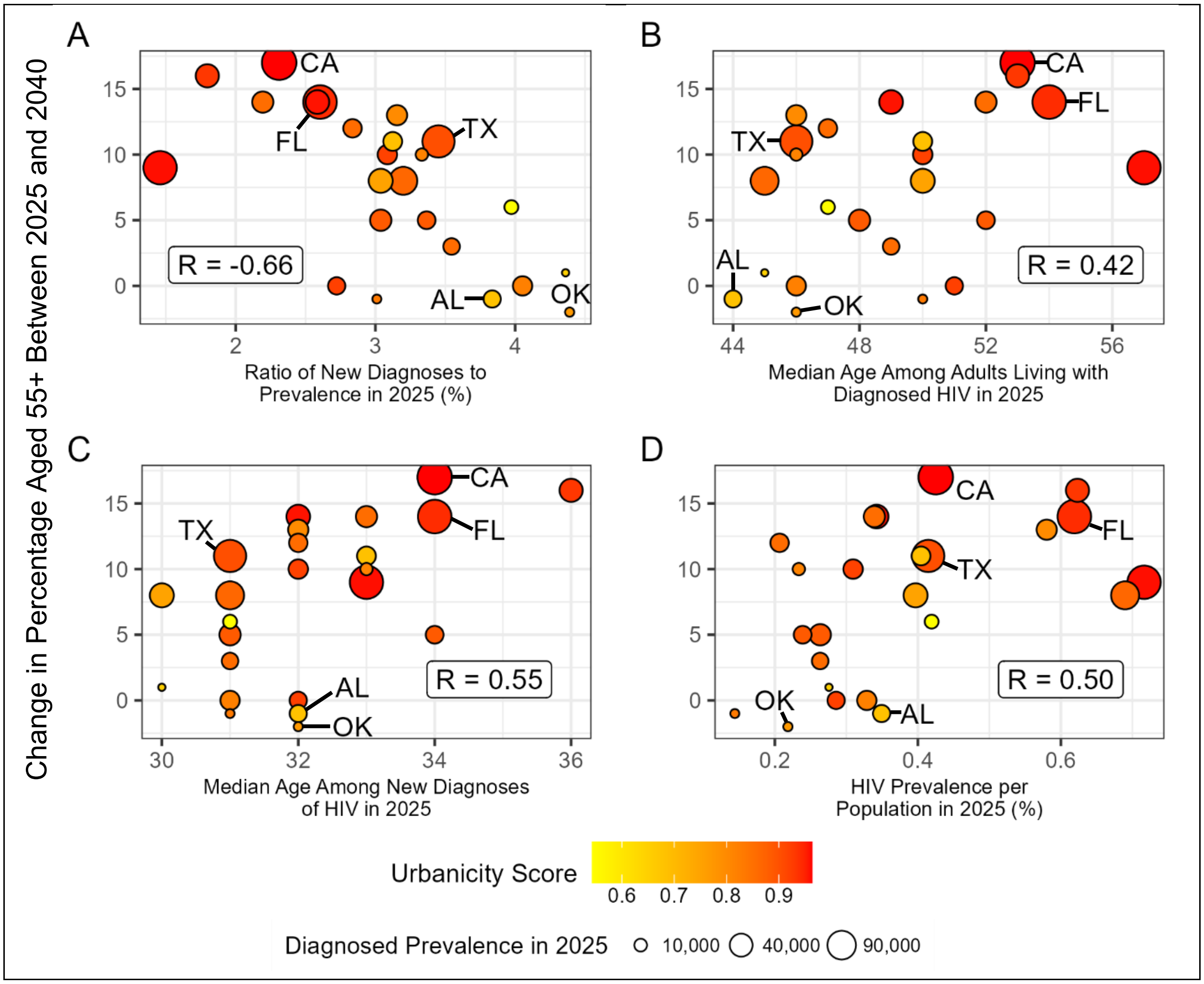

Scatterplots show the projected change in percentage of PWDH who are aged 55+ versus four state-level characteristics, averaged across 1,000 simulations in each state: (A) the ratio of new HIV diagnoses to HIV prevalence in 2025; (B) the median age among adults living with HIV in 2025; (C) the median age among new HIV diagnoses in 2025; and (D) prevalence of diagnosed HIV as percentage of the general population in 2025. Each point represents one of the 24 states; points are sized by that state's diagnosed prevalence in 2025 and are shaded by the urbanicity of the state's HIV epidemic, which has a Spearman correlation of 0.48 with the projected change in percentage of PWDH who are aged 55 or older. Five states are highlighted: Alabama ("AL"), California ("CA"), Florida ("FL"), Oklahoma ("OK"), and Texas ("TX"). Each panel is labeled with the Spearman correlation coefficient.

### Sensitivity Analysis

#### Supplemental Figure S6: Sensitivity analysis for parameters most strongly associated with change in median age of people with diagnosed HIV from 2025 to 2040

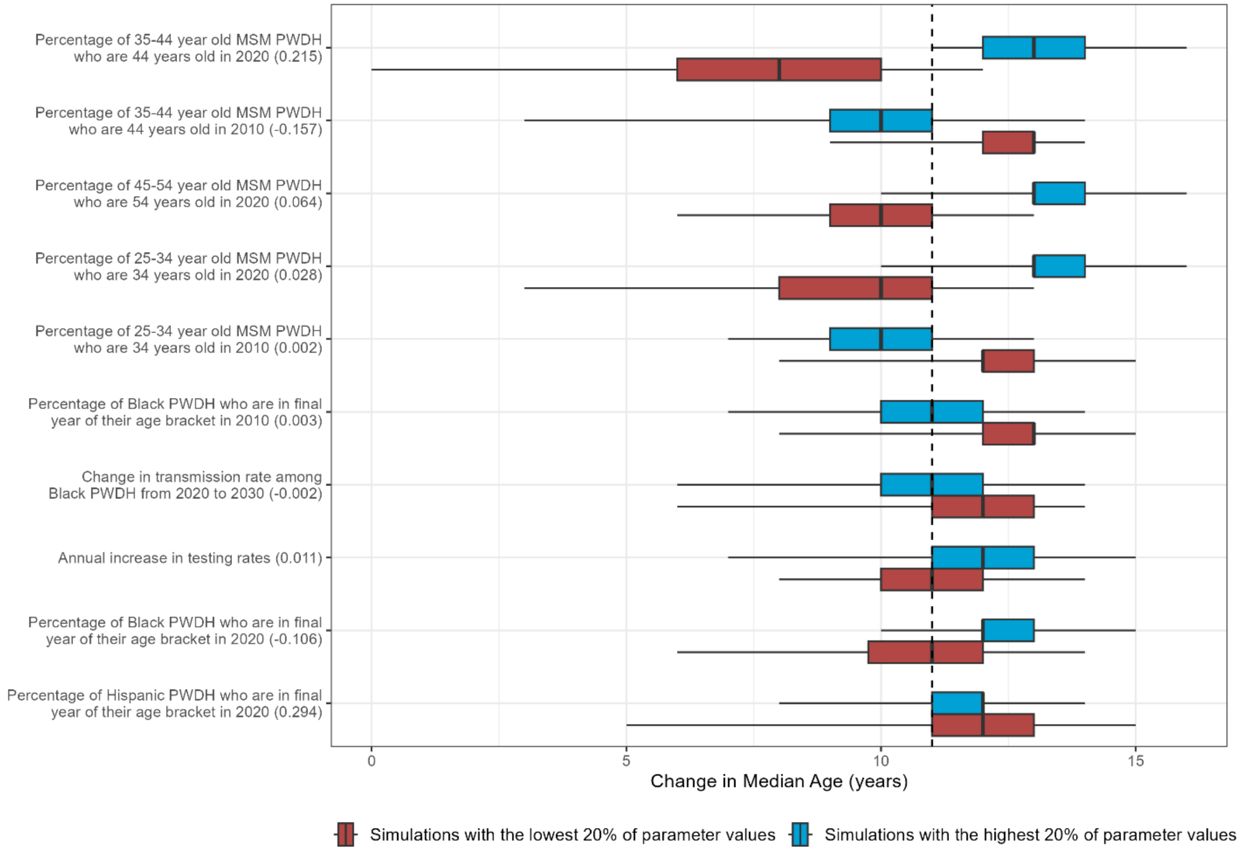

Boxplots show the projected change in median age from 2025 to 2040 across 24 states for the 200 simulations with the highest values of each parameter (blue) versus the 200 simulations with the lowest values (red). Black vertical lines give the median estimate across the 200 simulations; shaded boxes give the interquartile range; whiskers give the 95% credible interval. Parameters are ordered by the difference in median estimates between the two subsets. They are labeled with the average partial rank correlation coefficient across all states in parentheses. The dotted line shows the mean change in median age across the 24-state region, 11.

increase in cases vs. a scenario where Ryan White services continue uninterrupted. The dark vertical lines indicate the median projection across 1,000 simulations, the boxes indicate interquartile ranges (IQR), and whiskers cover the 95% credible interval. * = EHE priority states.
